# Neural systems underlying autobiographical memory dysregulations in depressive and at-risk individuals: A neuroimaging meta-analysis

**DOI:** 10.1101/2025.04.06.25325353

**Authors:** Cheuk Chi Charlotte Cheng, Hei Lam Michelle Tsang, Mengfan Han, Jie Zhang, Michael Maes, Benjamin Klugah-Brown, Mercy Chepngetich Bore, Benjamin Becker

## Abstract

**Background:** Autobiographical memory (AM) dysfunction has been proposed as a neurocognitive mechanism underlying the development and maintenance of depression. However, case-control neuroimaging studies investigating the neural correlates of AM in depression have yielded inconsistent findings. The present study utilized neuroimaging meta-analyses to identify robust neural markers of AM dysfunction in depression and to characterize the associated behavioural and network-level mechanisms.

**Methods:** A pre-registered neuroimaging meta-analysis (https://osf.io/35xtf) was conducted, incorporating data from 341 patients with unipolar depression and 261 healthy controls across case-control studies examining AM processing. Meta-analytic network-level and behavioural decoding analyses were performed to aid interpretation of the findings.

**Results:** Compared to controls, patients with depression showed increased activation in the right paracingulate cortex (dorsal anterior cingulate, dACC) and precuneus, and decreased activation in the anterior insula during AM recall. Exploratory valence-specific analyses revealed that negative AM recall was associated with increased activity the dACC and precuneus. Meta-analytic decoding linked the dACC to the salience network and to domains related to negative affect and executive control, while the precuneus was associated with the default mode network and with processes related to social cognition and autobiographical memory.

**Conclusions:** Findings do not support prevailing models emphasizing altered amygdala and hippocampal function in AM deficits in depression. Instead, they highlight the involvement of core regions within the salience and default mode networks as key neural substrates of AM dysfunction. These regions may contribute to affective, social-cognitive, and mnemonic disturbances that shape the valence-specific nature of AM deficits in depression.

## Introduction

Autobiographical memory (AM) is a memory system that consists of episodes recollected from an individual’s life, i.e. personal semantic and episodic information (Tulving 1972, 1983), with the former referring to facts about the self, such as information on where the individual was born, and the latter referring to unique events, such as the first time riding a bicycle (Brewer, 1996).

Dysfunctions such as impaired or biased AM recall have been observed across mood and anxiety disorders (Lenaert et al., 2015; Moscovitch et al., 2018; Weiss-Cowie et al., 2023), with accumulating evidence for an important role in the development and maintenance of unipolar depression (Weiss-Cowie et al., 2023). AM dysfunction in depressed individuals has been characterized by generally impaired recollection, as well as valence-specific effects, i.e. impaired memory for positive events in the context of a better memory for negative events (Burt et al., 1995; Matt et al., 1992). Within the context of cognitive models of depression such as Beck’s negative cognitive triad model (2008; Beck et al., 2024) proposing that negative beliefs about the self, the world, and the future shapes depression, a negative AM bias can subserve a vicious cycle of dysfunctional and self-enforcing negative schemas that contribute to the development and maintenance of depression (Beck et al., 2008, 2024.).

In experimental settings the behavioural and neural basis of AM and its alterations in mental disorders have been examined by combining the retrieval of general and emotional autobiographical memories, e.g. by cued recall, personalized scripts or prospection (St. Jacques & Cabeza, 2012) with functional Magnetic Resonance Imaging (fMRI, for details on the specific experimental paradigms see also Brewer, 1996, Young et al., 2012, 2013; Gilboa et al., 2004). Despite a certain level of inconsistency between the studies, recent meta-analyses confirm impairments in retrieving specific autobiographical memories in individuals with depression and increased risk for depression (Hallford et al., 2021; Hallford et al., 2022; Liu et al., 2013), with some original studies suggesting valence-specific effects, e.g. better memory for negative events (Kuyken & Dalgleish, 2011).

The neurobiological basis of AM has been examined in numerous fMRI studies suggesting that AM relies on circuits encompassing prefrontal regions as well as the hippocampal formation, retro splenial cortex and posterior cingulate cortex (Svoboda et al., 2006; Shephardson et al., 2023), with some evidence for differential involvement of the basal ganglia and amygdala/insula depending on the emotional content of the memories (e.g. Testa et al., 2024). The widespread engagement may reflect the complex interaction between mental processes during AM encompassing memory recall, self-referential processes, executive function, imagery and semantic contextualization. Within this network the hippocampal formation plays a pivotal role in the memory formation and recall (Fink et al., 1996; Gardini et al., 2006; Greenberg et al., 2005; Ryan et al., 2001) as well as in detailed and immersive recollection of memory details for AM, and is further supported by the medial prefrontal cortex (PFC) involved in the processing of self-referential stimuli (St. Jacques & Cabeza, 2012).

Furthermore, the lateral PFC, (specifically the left ventrolateral PFC), has been identified as a key region for memory search and retrieval in AM recall (Shepardson et al., 2023). Regions such as the amygdala or the basal ganglia have been identified to mediate the emotional content of AM, such that the amygdala has been involved in recalling negative events (Doré et al., 2018; McCrory et al., 2017) or the subjective sense of remembering visual (Greenberg & Rubin, 2003) and emotional (Rubin & Bernsten, 2003) re-experiencing of the event (Piolino et al., 2009). Affectively coloured AMs are evoked by stimulation of the amygdala (Vignal et al., 2007), and the activity of amygdala single neurons were associated with familiarity and recollection (Rutishauser et al., 2008), while the globus pallidus may mediate the emotional experience of positive AM (Testa et al., 2024).

More recent approaches examining the network level underpinnings of AM further indicate that the default mode network (DMN) - a large-scale brain network associated with self-referential processes, mind wandering and memory (Faustino, 2022; Philippi et al., 2015) plays a significant role in AM. A recent fMRI meta-analysis on AM by Shepardson et al. (2023) found that there is a large degree of overlap between the DMN and brain regions involved during AM retrieval, particularly in ‘core DMN’ regions, including frontal and posterior cortical midline structures and the bilateral angular gyrus (Andrews-Hannah et al., 2014; Shepardson et al., 2023).

While fMRI meta-analyses have allowed to robustly determine the neural systems underlying AM in healthy individuals (Shepardson et al., 2023) results on the neural systems mediating dysfunctional AM in depression are based on single studies that are characterized by a level of inconsistency inherent to case control fMRI studies (Etkin, 2019; Köhler et al., 2015; Klugah-Brown et al., 2021). The development of neuroimaging meta-analytic approaches facilitates quantitative integration of findings from case-control MRI studies, offering critical advancements towards determining more robust structural and functional alterations in depression within a behavioural domain or in comparison to other mental disorders (Bore et al., 2024b; Liu et al., 2022; Zhou et al., 2020).

Against this background, the present pre-registered neuroimaging meta-analysis aims to compensate for single-study deficits in aspects such as sample size and specific AM recall tasks that differ across studies. Neuroimaging meta-analyses have been developed to pool data extracted from original studies, and this allows for examination of brain activity across studies with a higher statistical power. As such a neuroimaging meta-analysis on AM recall in depressive and at-risk individuals can increase the robustness and generalisability of the results and allow to quantitatively determine common brain regions associated with AM recall in depressive individuals. Similar approaches have been recently applied to other domains and have indicated e.g. robust striatal alterations during reward processing in depression across domains and within at-risk populations (e.g. Lyu et al., 2024; Bore et al., 2024a, 2024b; Zhao et al., 2024) and may help to inform interventions that aim at targeting neural alterations in depression, e.g. by closed-loop real-time neuroimaging interventions (e.g. Young et al., 2017b; Misaki et al., 2024; Li et al., 2019).

Based on previous studies, we hypothesised that (1) neural alterations during AM in depression will be observed in the hippocampus, and amygdala, in particular decreased activity in the hippocampus, and increased amygdala response in depressive individuals when retrieving negative autobiographical memories (e.g. Doré et al., 2018; Young et al., 2016a; Young et al., 2016b), and that (2) the identified regions will connect on the network level with the DMN.

To test our hypotheses we capitalized on previous case-control neuroimaging studies on AM in depression and performed a coordinate-based meta-analysis on Seed-based d mapping with Permutation of Subject Images (SDM-PSI), a novel and robust meta-analytic technique that produces unbiased estimation of effect sizes and generation of neurofunctional maps (Albajes-Eizagirre et al., 2019), with subsequent meta-analytic co-activation and connectivity analyses determining the network level communication of the identified regions.

## Method

### Search strategy and selection criteria

The current pre-registered meta-analysis adhered to the guidelines of conducting a coordinate-based meta-analysis. A pre-registration was submitted on the Open Science Framework (OSF) (https://osf.io/35xtf) platform to increase accountability and transparency prior to the commencement of the meta-analysis. A comprehensive literature search was conducted independently according to the Preferred Reporting Items for Systematic Reviews and Meta-Analyses (PRISMA) guidelines by C.C.C.C. First, a broad search of original fMRI studies on fMRI case-control studies AM in depression was conducted on PubMed (https://pubmed.ncbi.nlm.nih.gov/) and Web of Science (https://www.webofscience.com/wos/). Suitable studies from the reference lists of review articles were additionally included. Literature was screened referring to the returned titles and abstracts. Studies that were written in English, reporting whole-brain results and were published between 1998-2023 were included. Only peer-reviewed, original case-control studies comparing patients and healthy controls were included. The following search items were applied: “functional magnetic resonance imaging OR fMRI” AND “AM” AND “depression OR major depressive disorder OR at-risk of depression” have to co-occur with any of the following keyword: “memory recall”. Data selection process was double checked by H.L.M.T and M.C.B, with any discrepancies settled by B.B. This search yielded 150 unique studies as shown in the PRISMA flow diagram (Page et al., 2021), see supplements.

Other exclusion criteria were: (a) functional connectivity, region of interest (ROI) and small-volume correction (SVC) studies, (b) studies on participants <18 years of age, (c) resting state fMRI studies, (d) studies focusing solely on other mental disorders, including bipolar disorder, posttraumatic stress disorder, and borderline personality disorder, (e) studies that reported clusters from the same participant pool of already included studies. The systematic literature review identified 13 suitable studies.

### Coordinate-based meta-analytic approach

A coordinate based meta-analysis of AM in individuals with or at risk of depression was conducted using Seed-based d Mapping Permutation of Subject Images (SDM-PSI) (https://www.sdmproject.com) which performs statistical inferences on results from multiple studies (Albajes-Eizagirre et al., 2019) and has been successfully employed to determine robust neurofunctional alterations within or across mental disorders (see e.g. Bore et al., 2024b; He et al., 2025). Peak coordinates and effect sizes of case-control differences during AM were extracted from the original studies reporting results on the whole-brain level.

Our major aim was to identify brain functional alterations during AM recall and its directionality in depression by implementing: (a) extraction of peak coordinates of between-group differences (depressive or at-risk individuals versus healthy subjects) and their effect sizes (p values or t/z values), (b) estimation of the lower and upper bounds (Hedges’ g) of the most probable effect sizes, inter-study heterogeneity was examined by evaluating the *I*^2^ index of the main clusters; (c) Meta-analysis of Studies with Non-Statistically Significant Unreported Effects (MetaNSUE) to estimate the probable effect size and standard error, as well as create imputations with reference to the estimations that lie within the bounds; (d) definition of the study specific alteration map between depressive or at-risk subject individuals and controls; (e) permutation test of subject images to allow statistical evidence (Jauhar et al., 2021). Statistical significance testing with a threshold of voxel-level p ≤ .0025 uncorrected was applied in order to balance statistical errors of Type 1 and 2 as shown in previous studies (Bore et al., 2024b; Liu et al., 2022). The SDM algorithm adopted an anisotropic Gaussian kernel with FWHM = 20mm, voxel size = 2mm.

Z-values representing between-group differences of depressed or at-risk individuals and healthy controls were transformed to t-values using the SDM statistical converter (https://www.sdmproject.com/utilities/?show=Coordinates). MNI coordinates (x, y, z) of activation clusters with statistically significant differences across tested contrasts will be reported.

Supplementary analyses were conducted on Neurosynth to explore the functional connectivity and meta-analytic co-activation patterns of the identified regions. Functional connectivity reflects brain regions that are coactivated across the resting-state fMRI time series with the seed voxel (Yeo et al., 2011) while meta-analytic coactivation employs all of the fMRI studies available in the Neurosynth database and serves as the meta-analytic analogue of the functional connectivity map (Rottschy et al. 2013, Schnellbächer et al. 2020).

### Analyses of the effects of valence AM recall

Given the frequently reported valence-specific effects in depression (Liu et al., 2021; Köhler et al., 2015), we conducted further exploratory analyses (Whalley et al., 2012, Wu et al., 2020). To identify possible effects of valence AM on brain region activation in AM recall, three sub analyses were performed based on evidence from the literature on valence effects on brain activity – one with all types of valenced AM recall, one with positive AM recall, and one with negative AM recall (McCrory et al., 2017; Young 2012, 2013, 2014). Specifically, the neural mechanisms may differ between positive and negative AM recall among healthy controls and depressed participants. The sub analysis for valence AM excluded some statistically significant data points from 5 studies (Young et al., 2012, 2013, 2014, 2015, 2016b); the analysis of positive AM included 9 studies, and the analysis of negative AM included 7 studies. Due to the low number of studies included in the sub analysis, findings should be considered as exploratory, and would need a more extensive investigation of valence effects.

### Analyses of the effects of depression in AM recall

As high-risk participants may not have the same neural deficits to depressed patients, a sub analysis with only participants with depression and remitted depression was carried out. This was to identify possible effects of depression on AM on brain region activation in AM recall, and to simultaneously control possible biases from individuals not matching the full criteria for depression. This led to a sub analysis that included 12 studies, with one study being fully excluded (MacDonald et al., 2016) and some datapoints from 2 studies were further excluded (Young et al., 2013, 2015).

### Additional analyses

Using SDM values and other confounding variables such as age and sex, we performed meta-regression analyses.

### Tests for heterogeneity and publication bias

Heterogeneity and publication bias tests were performed with standard procedures. Tests for homogeneity are important to consider studies in meta-analyses that used similar experimental protocols and procedures. The I^2^ index, which is a representation of the proportion of total variation in heterogeneity derived from studies, was used to examine between-study heterogeneity for all significant clusters of the main meta-analyses (Higgins and Thompson, 2002). I^2^ scores indicate low (>25%), moderate (>50%) and high (>70%) heterogeneity, in which high heterogeneity may indicate reduced generalisability of results (Martins et al., 2021). Publication bias was statistically evaluated by Egger’s test and funnel plots, where p values <.05 were interpreted as significant.

## Results

### Demographic and clinical data summary of all included studies

A total of 13 studies comprising of 602 participants (*n* = 261 healthy controls, *n* = 341 patients) were included in the meta-analysis. There were 12 unique participant pools identified, as Young (2013, 2016a) used the same participants in two separate studies. With exception of some studies which did not provide sufficient information (Gillard et al., 2023; MacDonald, 2016; Whalley et al., 2012; Wu et al., 2020), participants had a mean age of 34.41 (*SD* = 10.761). Healthy controls (*n* = 156) had a mean age of 34.17 (*SD* = 11.169), and depressive participants (*n* = 384) (including high risk, depressive and remitted depressive patients) had a mean age of 34.52 (*SD* = 10.604). There were no significant group differences between healthy controls and patients (*t* = .342, *p* = .732). The prism flow diagram as well as a complete list of included studies and their design characteristics are shown in **Fig. 1** and **Table 1**.

**Figure 1.**
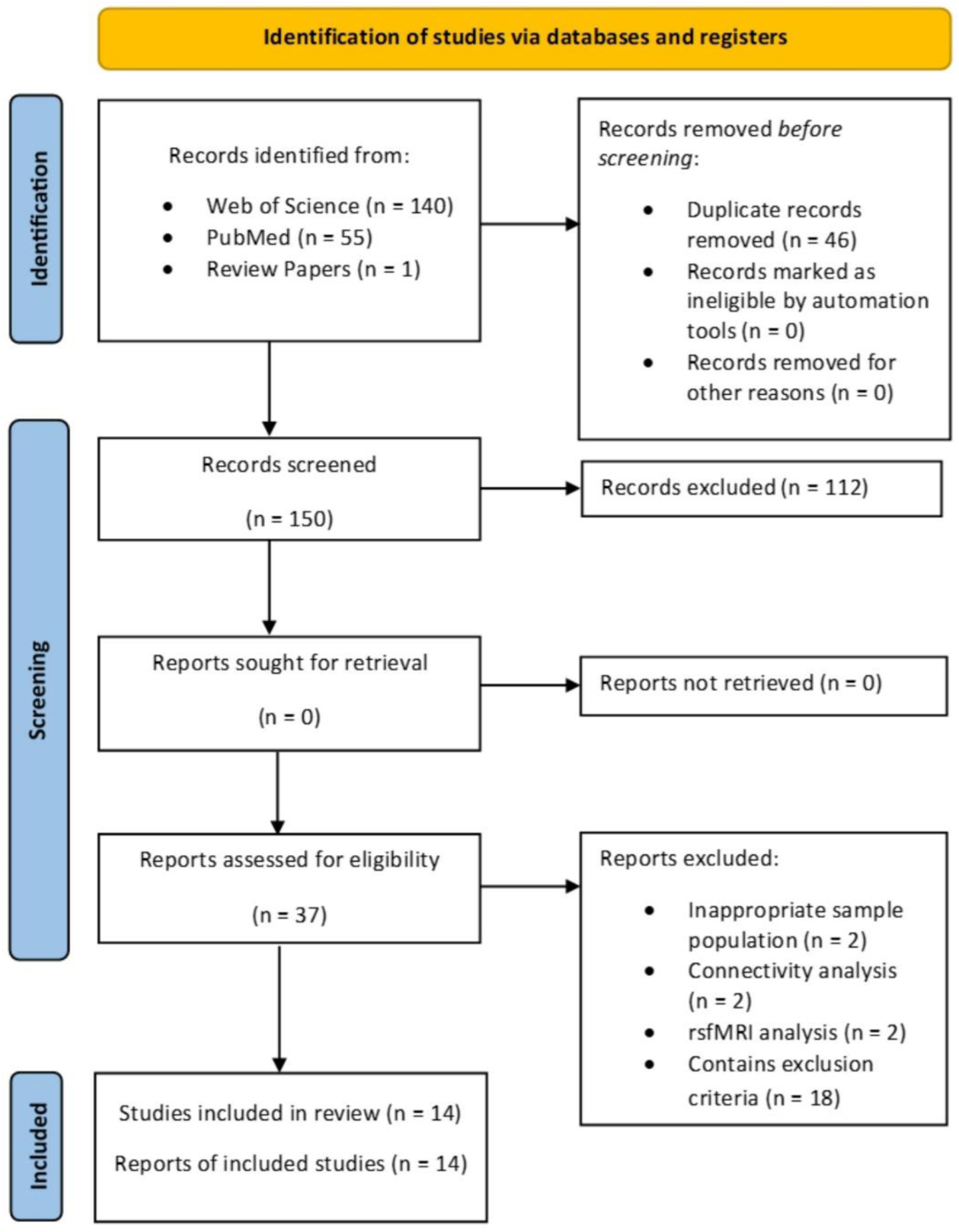
PRISMA flow diagram showing the identification of included studies

**Table 1.** Demographic and clinical characteristics of included studies.

| Study | N |  |  |  | Control Task | Active Task |
| --- | --- | --- | --- | --- | --- | --- |
|  | HC | MDD | rMDD | HR |  |  |
| Foland-Ross et al., 2014 | 16 | 0 | 16 | 0 | Centre cross fixation and sad mood inducement | Cue words - novel (positive) |
| Gillard et al., 2023 | 21 | 18 | 0 | 0 | 30 seconds closed-eye rest | Pre-recorded voice recording of their AM memories (neutral, social inclusion and social rejection) |
| Keedwell et al., 2005 | 12 | 12 | 0 | 0 | Happy, sad and neutral facial expressions | Pre-recorded voice recording of their AM memories |
| MacDonald et al., 2016 | 21 | 0 | 0 | 16 | 30 seconds rest | Cue sentences - rehearsed |
| Parlar et al.,<br>2018 | 12 | 12 | 0 | 0 | Odd number detection task | Visual cues with cue words - novel |
| Whalley et<br>al., 2012 | 15 | 15 | 0 | 0 | Cue words and sentences -<br>rehearsed (identify if it is from<br>someone else's narrative) | Cue words and sentences - rehearsed (identify if<br>it is from their own narrative) |
| Wu et al.,<br>2020 | 18 | 16 | 0 | 0 | Visual neutral photos displayed | Visual cues – novel |
| Young et al.,<br>2012 | 14 | 12 | 0 | 0 | Subtraction task | Cue words - novel (positive, negative and<br>neutral) |
| Young et al.,<br>2013, 2016a | 16 | 16 | 0 | 16 | Semantic example generation<br>tasks from positive, negative<br>and neutral cue words and riser<br>detection task for non-word<br>letter strings | Cue words - novel (positive, negative and<br>neutral) |
| Young et al.,<br>2014 | 16 | 16 | 16 | 0 | Semantic example generation<br>tasks from positive, negative<br>and neutral cue words | Cue words - novel (positive, negative and<br>neutral) |
| Young et al.,<br>2015 | 20 | 0 | 20 | 20 | Semantic example generation<br>tasks from positive, negative<br>and neutral cue words and riser<br>detection task for non-word<br>letter strings | Cue words - novel (positive, negative and<br>neutral) |
| Young et al.,<br>2016b | 60 | 45 | 25 | 30 | Riser detection task for non-word letter strings | Cue words - novel (positive, negative and<br>neutral) |
| Young et al.,<br>2017 | 40 | 40 | 0 | 0 | Semantic example generation<br>tasks from positive, negative<br>and neutral cue words | Cue words - novel (positive, negative and<br>neutral) |

Abbreviations: HC, healthy control; MDD, major depressive disorder; rMDD, remitted major depression disorder; HR, high-risk

### Main meta-analytic findings

The overall meta-analysis encompassing both positive and negative valence AM recall revealed three significant clusters of increased activation in depression relative to controls located in the right anterior cingulate/paracingulate, right precuneus and the right middle temporal gyrus, and a single cluster of decreased activity in the right anterior insula. Sub-group meta-analyses investigating the effects of negative and positive valence AM recall further demonstrated that patients exhibited increased activity in the cingulate/paracingulate cingulate and right precuneus during negative recall but increased activity in the right inferior temporal gyrus during positive recall (**Fig. 2**, **Table 2**).

**Figure 2.**
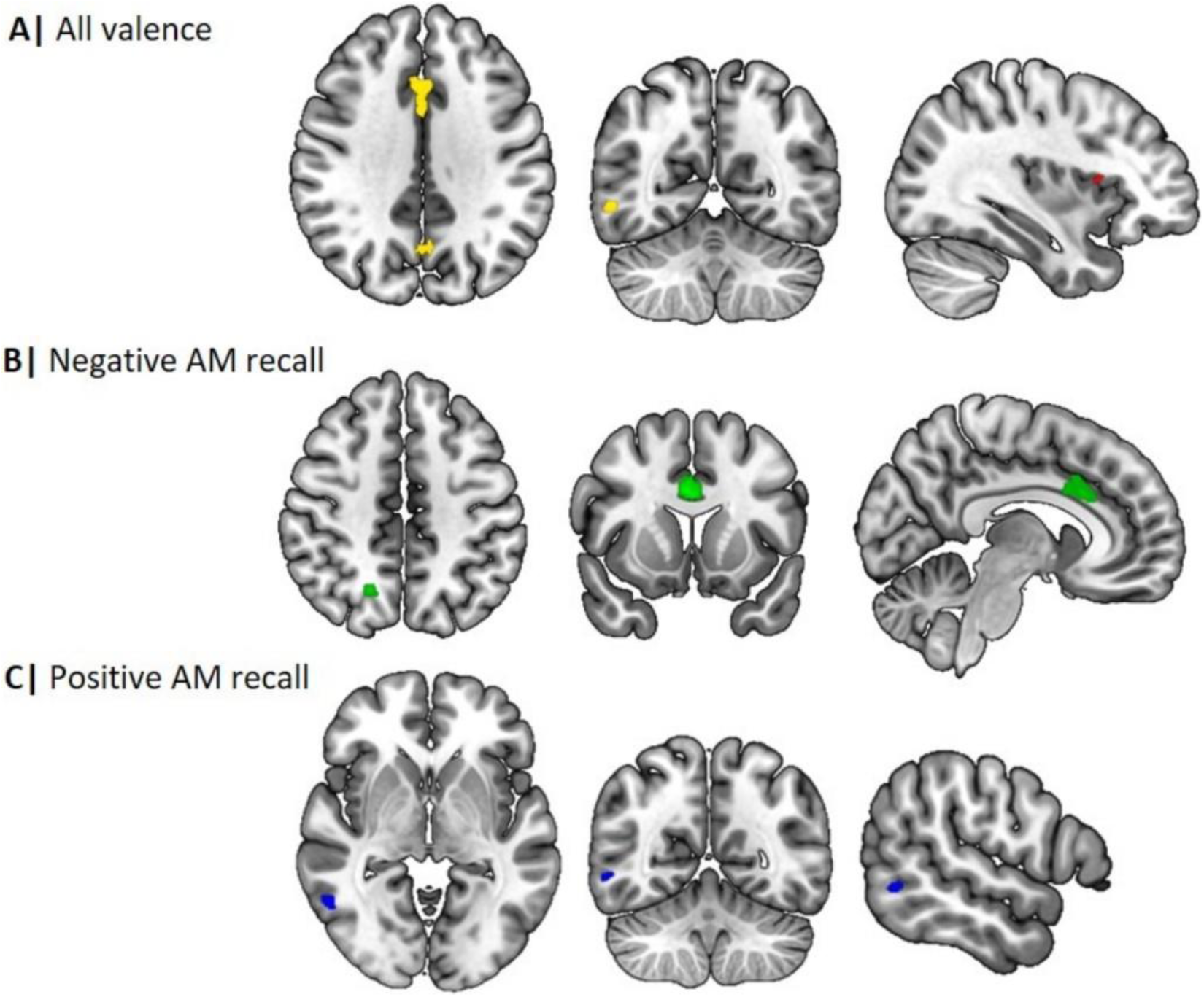
Illustration of the main meta-analytic findings

**Table 2.**
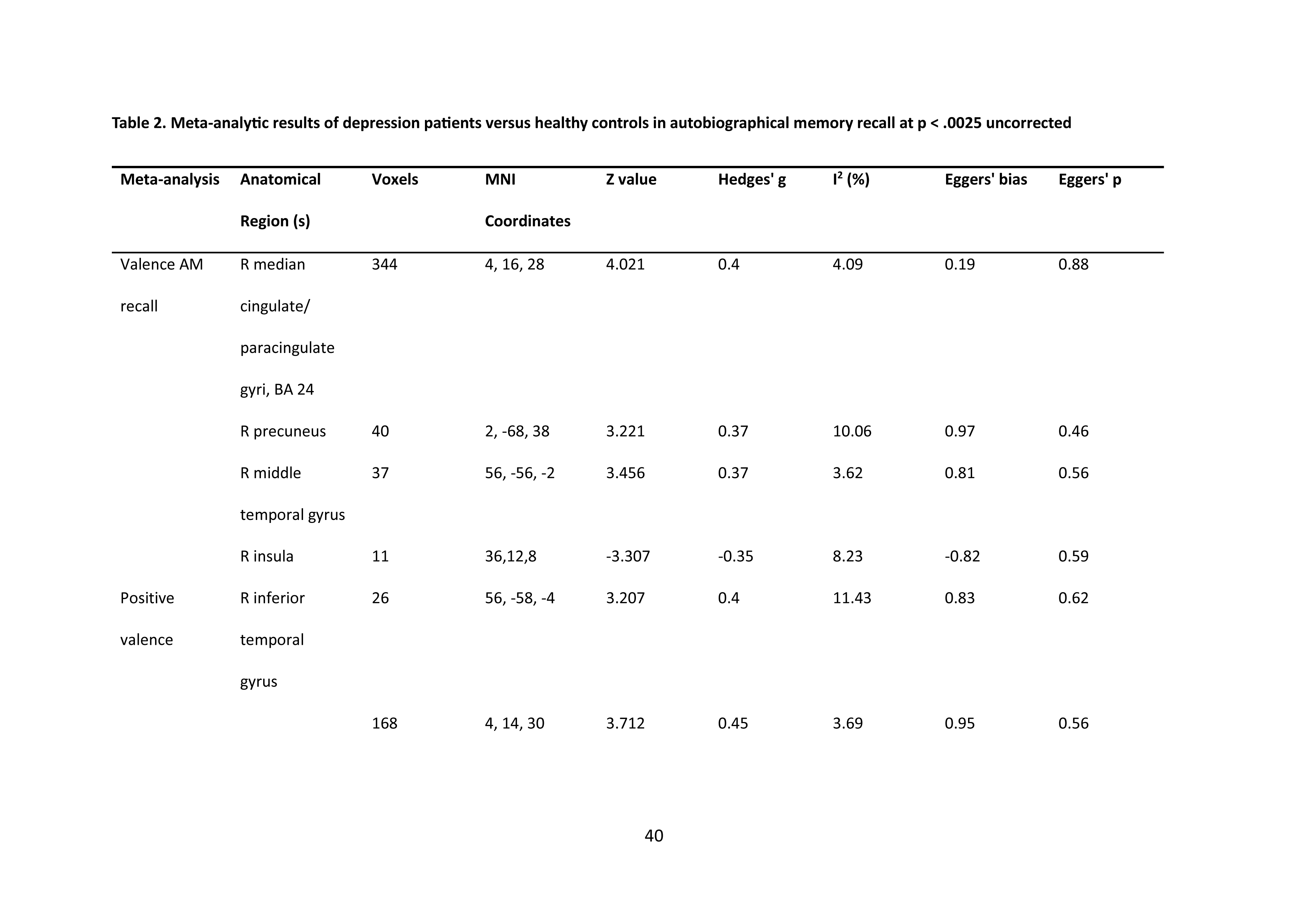
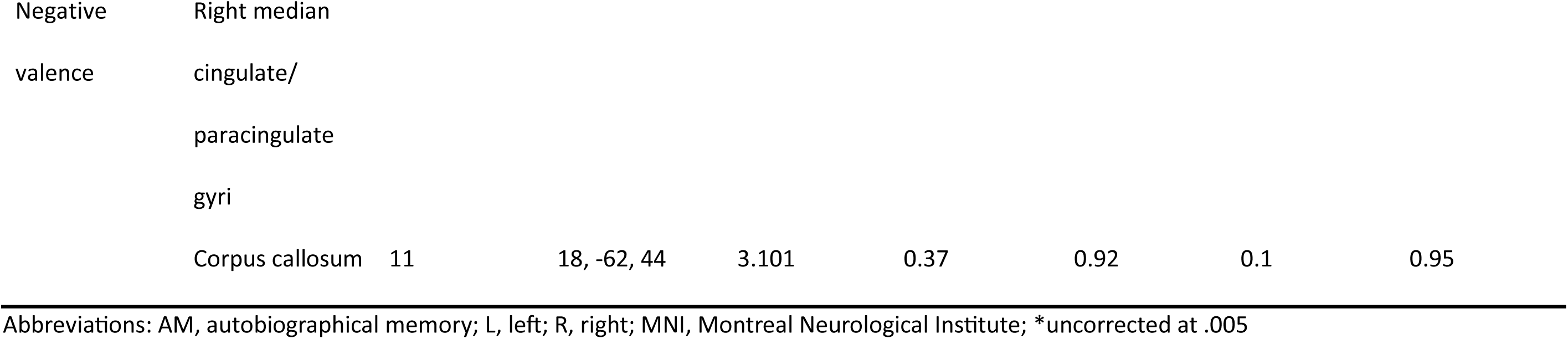
Meta-analytic results of depression patients versus healthy controls in autobiographical memory recall at p < .0025 uncorrected.

### Functional connectivity and behavioural level analyses

Additional analyses were conducted on the network and behavioural levels. Network analyses were performed utilizing the Neurosynth database (https://www.neurosynth.org/) to explore the functional connectivity and meta-analytic coactivation patterns of the paracingulate and precuneus regions identified in the general valence meta-analysis as shown in **Fig. 3A** and **Fig. 3B** respectively.

**Figure 3.**
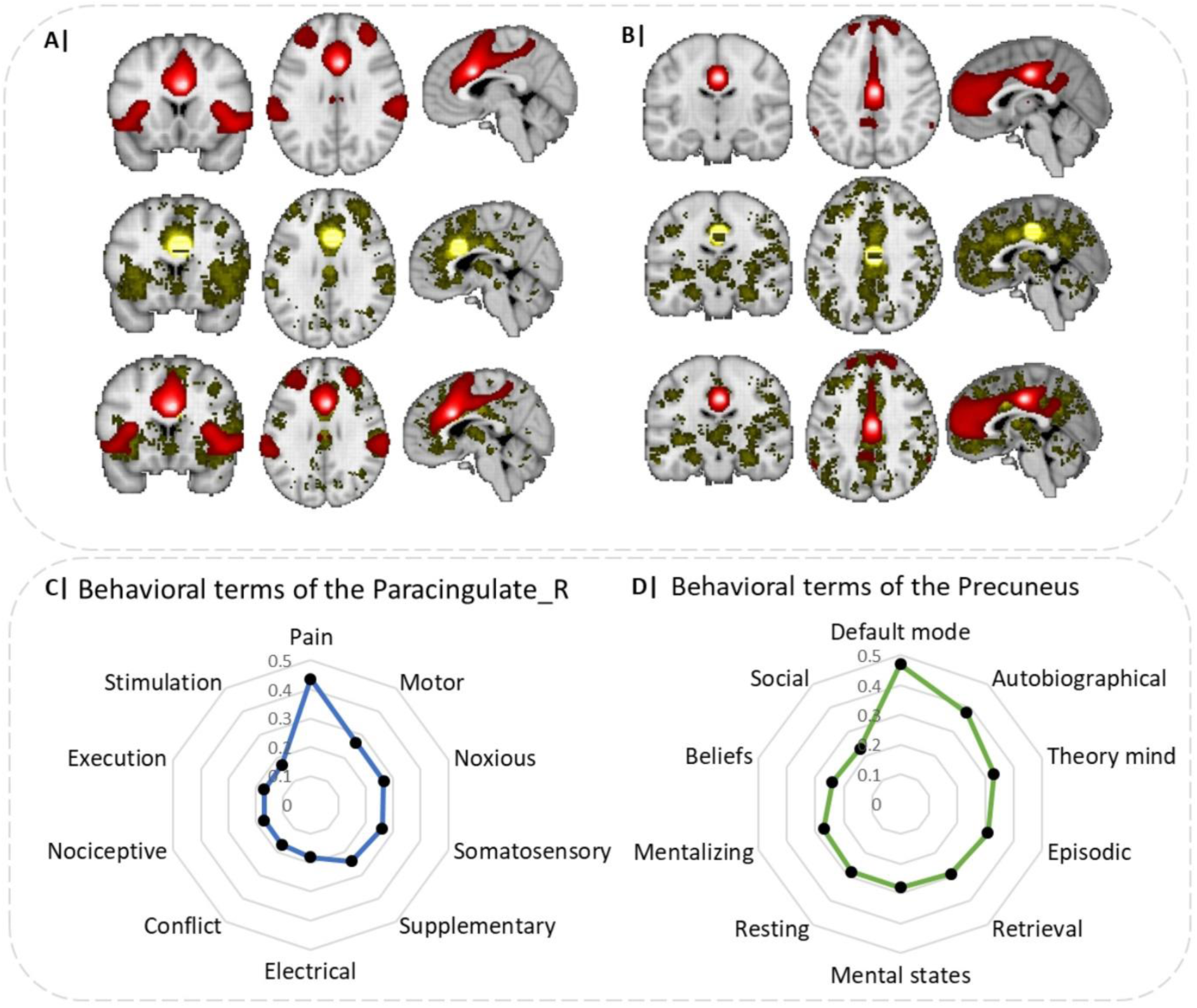
A and B Functional connectivity and meta-analytic co-activation of the regions identified of being altered in depression during AM recall for paracingulate regions, and precuneus respectively. Top panel shows the functional connectivity, middle panel shows the meta-analytic co-activation patterns and bottom panel shows the combination of both functional and meta-analytic co-activation patterns. Behavioral terms of the paracingulate and precuneus are shown in C and D respectively.

The top panel shows the strong functional connectivity of the paracingulate with core regions of the salience network, including the ACC as well as the bilateral anterior insula (extending into the inferior frontal gyrus). Overlap of the functional connectivity and meta-analytic coactivation maps confirmed that core connectivity profiles encompassed the anterior insula and anterior cingulate cortex (**Fig. 3A**). The functional connectivity for the seed voxel in the precuneus showed interactions with core cortical midline regions of the default mode network, in particular medial frontal regions as well as the precuneus and PCC. Overlap of the functional connectivity and meta-analytic coactivation maps confirmed functional interactions with the cortical midline regions (**Fig. 3B**).

The additional behavioral characterization indicated that the paracingulate region is strongly associated with negative affective (e.g. pain) and executive (conflict) functions, while the precuneus regions is associated with social cognitive and mnemonic functions, including autobiographic and theory (of) mind as shown in **Fig. 3C and Fig. 3D** respectively.

### Additional analyses: Meta-regression

Meta-regression analyses of age and gender as possible confounding variables were conducted. The analysis found no significant associations as shown on **Fig. S1**.

### Tests for Heterogeneity and Publication bias

Between-study heterogeneity was generally low across the three analyses. With reference to the bias p-values and symmetric funnel plots, publication bias analysis was not statistically significant as shown by smaller p-values in the Egger’s p column (see Table 2 for more information).

## Discussion

Our main meta-analysis identified increased activation in the right median cingulate/paracingulate gyri (overlapping with the dorsal ACC), left precuneus and right inferior temporal gyrus and concomitantly increased activity in the right anterior insula in depressive individuals compared to healthy controls during autobiographical memory recall. Moreover, exploratory valence-specific meta-analyses revealed increased activity in an overlapping network during negative recall (i.e. dorsal ACC and precuneus) and positive recall (inferior temporal gyrus) suggesting potentially valence-specific effects. On the network level, the core regions encompassing the dorsal ACC and precuneus exhibited functional interactions with the salience network and the DMN, respectively. Behavioural characterization further revealed an engagement in negative affect and executive functions of the dorsal ACC and autobiographical memory and social cognition of the identifies precuneus region respectively.

While the functional and behavioural characterization aligns with the corresponding deficits in these domains in depression (e.g. Xu et al., 2022; Young et al., 2012) the identified regions did not align with our hypothesized changes including reduced activation of the hippocampus, and increased activation of the amygdala during AM in depression. The dACC is not considered to play a key role in AM per se and in the context of mnemonic functions has been associated with remote AM recall, which together with a recent meta-analysis suggests a potential contribution to AM (Daviddi et al.,2023; Steinvorth et al., 2006) (see also Martinelli et al., 2013). However, in line with the meta-analytic functional and network level characterization the dACC plays a key role in affective and executive functions, including conflict processing and negative emotional appraisal (Etkin et al., 2011; Li et al., 2020), with recent work from Gillard et al. (2023) further suggesting that the dACC could be linked to activations across the affective salience network as a response to the psychological experience of social pain and affiliation (Dalgleish et al., 2017; Eisenberger et al., 2003), which aligns with previous findings indicating altered activity in this region during negative AM recall in depression (Young et al., 2017a).

Previous studies on dACC in depression have reported robust alterations in the functional intrinsic network architecture of this region (Zhou et al., 2024) and an association with different symptom domains in depression (Sheline et al., 2010). During task engagement this region has demonstrated altered functional communication during social cognitive processes, including e.g. social working memory ( Xu et al., 2022), which together with the stronger contribution of this region in the negative AM studies on depression may reflect that dysregulations in this area may contribute to an overgeneralized memory in depression (Falco et al., 2015; Raes et al., 2023) or mediate affective emotional aspects as well as general cognitive deficits in depression leading to emotionally-biased re-experience of social affective experiences (Gillard et al., 2023).

The precuneus has been strongly associated with AM retrieval, with research on the specific functions indicating a contribution to autobiographical reminiscence (Daviddi et al., 2023; Shepardson et al., 2023; Sreekumar et al., 2018), or first person visual and memory-based navigation (Wilson et al., 2005). It further shows its association with self-referential processing and self-consciousness (Freton et al., 2014). Interestingly, Freton et al. (2014) found that the spontaneous tendency to recall AM is positively correlated to precuneus volume which may imply a contribution to spontaneous AM retrieval. Within recent pathological models of negative emotional experiences, e.g. in PTSD, posteromedial regions such as the precuneus have been related to dysregulations in visual imagery processes (Thome et al., 2020). The contribution of this region to autobiographical, self-referential processing and social cognitive functions is further underscored by or behavioural characterization and the meta-analytic characterization as central part of the default mode network (see also Xin et al., 2021). Together the findings may reflect that alterations in this region may mediate the mnemonic deficits in the patients, potentially reflecting overgeneralized memory recall or dysfunctional imaginary and self-referential processes in depression. Decreased spontaneous activity in the precuneus has been demonstrated in depression – yet not related disorders such as bipolar disorder (e.g. Gong et al., 2020) – with recent evidence indicating a role of altered precuneus engagement during aberrant self-referential AM processing adolescents with depression (van Houtum et al., 2023) as well as less vividness in AM retrieval in depression (van Schie et al., 2019). This further supports the notion that depressed individuals are less likely to retrieve vivid AM and may do so in the context of aberrant self-referential appraisal.

The third significant cluster of increased activation was in the inferior temporal gyrus, a subregion of the DMN. Although the specific role of the inferior temporal gyrus is not well explored, Sheline et al. (2009) proposed that depressed individuals do not inhibit DMN activity while looking at negative pictures, which could possibly be linked to negatively valenced material. Lemogne et al. (2012) further supported this finding by indicating that depressive self-focus was related to the lack of DMN inhibition. This implies that there are underlying mechanisms regarding depressive individuals’ possible fixation on negative AM contributing to the maintainence of depressive symptoms.

Conversely, the SDM-PSI analysis found reduced activation of the right anterior insula. The insula is important in supporting subjective feeling states or interoceptive processes (Critchley et al., 2004; Zhang et al., 2023) and more vivid memories are associated with the activation of the insula (van Schie et al., 2019). With this in mind, it could be proposed that depressive patients are more likely to have reduced vividness in AM recall, which is indicative of decreased awareness of oneself. This implies that depressive patients are more distant when recalling AM, which is supported by Beck (1979), who posits that distancing oneself from a dissonant past self, or evaluating the self with a distance, are processes that are central to proposed theoretical accounts of depression. This was also supported by a study led by Kuyken and Howell (2006), which found that depressive patients have a more distanced view of their AM as compared to healthy individuals. The decreased activation of the insula could further suggest that depressive patients are more likely to have lowered awareness of the self, and an affected AM recall.

Functional connectivity analysis of the seed voxel in the paracingulate identified associations with the brain regions in the salience network, including the anterior insula, and the anterior cingulate cortex (ACC; Uddin et al., 2019). The anterior insula plays an important role in supporting both, subjective feeling states of emotional experience and autonomic reactivity (Ferraro et al., 2022) which may reflect that depressive individuals are more likely to fixate on negative emotional and autonomic experiences of AM (Dillon & Pizzagalli, 2018). In line with this interpretation previous studies indicate an engagement of the insula during recollection or reflection on personal distress (Carr et al., 2003; Wager et al., 2003) and evaluation of negative emotional states (Sanfey et al., 2003). Associations with the anterior cingulate cortex supports previous research proposing that AM retrieval relates to subjectively painful experiences (Kelly et al., 2007; Dalgleish et al., 2017; Eisenberger et al., 2003). Together, the involvement of the salience network may reflect a more aversive affective experience during AM in depression or a dysregulated attentional balance between internally and externally focused attention (Gillard et al., 2023; Xin et al., 2021).

Furthermore, the functional connectivity analysis for the seed voxel in the precuneus points to correlations to the DMN. The DMN, more specifically the anterior medial prefrontal cortex (amPFC) and the posterior cingulate cortex (PCC), is crucial in AM retrieval as it facilitates self-referential processing (Andrews-Hanna et al., 2014). Individuals with MDD, or individuals with high risk of developing depression show alterations in the activation of DMN (Chou et al., 2023; Posner et al., 2016). Depressed individuals tend to show differential engagement in the DMN and self-referential processes during task and rest conditions (Chou et al., 2023). Due to its core function and possibly causal contribution to free but not cued AM (Bonnici et al., 2018; Lanius et al., 2020), alterations in the DMN may reflect impaired free AM recall in depression. Finally, the corresponding behavioral characterization supports the engagement of both large-scale networks with the large-scale networks and suggests that dysbalanced engagement of the salience network and DMN facilitate AM deficits in depression.

Findings need to be considered in the context of limitations. Firstly, several studies utilise specific AM recall (Young et al., 2012, 2013, 2014, 2015, 2016a, 2016b), posing difficulties in determining the valence of AM retrieved. While we aimed at disentangling valence-specific effects utilization of the valence-specific results should be interpreted with caution due to the comparably low number of studies (Dahlgren et al., 2020; Ding et al., 2020). Secondly, there has been increasing support of semantic and episodic AM retrieval having independent neural networks and brain regions involved (Maguire et al., 2000; Levine et al., 2004) and with an increasing number of studies meta-analyses can focus on depression-related dysfunctions in episodic and semantic AM components. The studies were mostly conducted in western populations (exception: Wu et al. (2020) with participants from Southwest China). Studies have proposed that cultural contexts modulate the development of episodic AM (Harbus, 2010) and an increasing number of studies employs a cross-cultural neuroimaging approach (Yang et al., 2023).

In summary, the present meta-analysis does not support prevailing models that suggest amygdala and hippocampal engagement underlying autobiographical memory deficits in depression. Rather, results support the role of core nodes in the salience (dorsal ACC) and default mode network (precuneus) as a pathological substrate of AM dysfunctions among people with depression or those at risk. With the dACC being linked to affective and executive functions, results suggest that its activation could be associated with activations in the salience network as a response to the psychological experience of social pain and affiliation. Findings indicate that activation in the precuneus is in line with current research, further emphasising its role in AM retrieval, specifically in overgeneralised memory recall or dysfunctional imaginary and self-referential processes in depression.

## Funding sources

The study was supported by Ministry of Science and Technology, MOST2030, Grant No. 2022ZD0208500 and a start-up grant from The University of Hong Kong.

## Conflict of interest

The authors declare no conflict of interest.

## Data availability

All included studies have been cited within the paper. Data supporting the results of this paper are available at the Open Science Repository platform (OSF) (https://osf.io/35xtf/).

## Notes

### Competing Interest Statement

The authors have declared no competing interest.

